# Customization and acceptability of the WHO Labor care guide to improve labor monitoring among health workers in Uganda. An Iterative Development Study

**DOI:** 10.1101/2024.01.07.24300952

**Authors:** Mugyenyi R Godfrey, Byamugisha K Josaphat, Tumuhimbise Wilson, Atukunda C Esther, Yarine T Fajardo

**Author notes:** **Corresponding author:** Mugyenyi R Godfrey, Tele Department of Obstetrics and Gynaecology, Mbarara University of Science and Technology.

## Abstract

**Background:** Cognisant of the persistent maternal and perinatal mortality rates, the WHO has called for adoption and evaluation of new adaptable and context-specific health solutions to improve labor monitoring and health outcomes. We aimed at customizing and refining the new WHO labour care guide (LCG) to suite health care provider (HCP) needs in monitoring labour in Southwestern Uganda.

**Methods:** We used an iterative approach to customize and refine the new WHO LCG. Between 1^st^ July 2023 and 30^th^ November 2023, we conducted; 1)30 stakeholder interviews to identify user needs and challenges, and inform the first LCG modifications; 2)10 HCP exit interviews to obtain feedback and modify LCG prototype one; 3)Two focus group discussions following use of prototype two to identify any further user needs; 4)Exit expert panel interviews involving HCPs to refine LCG components; 5)Pilot testing of final prototype among 40 HCPs; 6)Final panel reviews from two expert conferences, the National Safe Motherhood Conference, and Association of Obstetricians and Gynaecologists of Uganda to refine/consolidate modifications of final prototype for Uganda, ready for evaluation.

**Results:** A total of 120 HCPs and MOH officials previously exposed to the new WHO LCG, with median age of 36 years (IQR;26-48) were interviewed. Over 53 modifications were made to tailor the WHO LCG into the modified LCG prototype for Uganda including; 1)Adjusting observation ordering to improve flow, clarity, and facilitate an easy user interface; 2)Inclusion of vital socio-demographic data compatible with existing programs in Uganda to prompt risk identification; 3)Modification of medications, baby-mother parameters/observations to suit local context; and 4)Inclusion of key cues to action, clinical notes and labour outcome data to facilitate auditing, accountability, reference, utilization and immediate postpartum care. All HCPs found the modified LCG useful, easy to use, appropriate, comprehensive, inclusive and would recommend it to others for use. Over 80% HCPs reported they took <2 minutes to plot/fill all observations on the LCG after assessment.

**Conclusions:** Active involvement of targeted end-users in customizing the LCG was observed to improve inclusiveness, ownership, comprehensiveness, acceptability, engagement and uptake. The modified LCG prototype was found to be simple, appropriate and easy-to-use. Further research to evaluate this LCG prototype feasibility and effectiveness is needed.

## Introduction

Uganda’s maternal mortality rate remains unacceptably high, and one of the highest in the world at 336 per 100,000 live births; perinatal mortality rates are 41/1000 live births [1]. To avert the persistent maternal and perinatal deaths, scholars have discouraged use of ineffective practices in labor and childbirth [2]; WHO has called for adoption and evaluation of new adaptable and context-specific health solutions to improve labor monitoring and promote better health outcomes [3, 4]. There has been difficulty in drawing useful conclusions about costs, and adaptability of new approaches aimed at improving health outcomes because of complexity and methodologies used to integrate new health interventions [5, 6]. However, end-user centered iterative approaches that aim at characterizing, adopting, refining and integrating the new approaches into routine care have been observed to improve uptake and sustained use of these interventions among intended users [7].

For over 5 decades, a partograph has been the standard tool used to monitor labour globally. However, despite decades of training and investment in using this tool for labour monitoring, the rates of maternal-perinatal outcomes remain poor in many countries. Some scholars have argued that the partogram majorly offers subjective variations, and assumes that all women progress at same rate, which may affect intervention rate and affect health outcomes for both mother and baby [8]. This subjective nature, coupled with the reportedly low acceptability of this partograph have grossly affected effective use of the partograph to monitor labour among others [8, 9]. Cognisant of the persistent maternal and perinatal mortality rates, the World Health Organization (WHO) has made new recommendations on intrapartum care to improve labour monitoring as well as maternal childbirth experience [4, 10]. The WHO has recommended that the partograph, a tool traditionally used to monitor labour globally, be modified to fit emerging user needs, available evidence and global priorities aimed at reducing the persistently high maternal-perinatal mortalities and morbidities [11–13].

WHO guidelines and recommendations included new definitions and considerations for durations of the first and second stages of labour, highlighted the importance of incorporating woman-centred care within the monitoring tool to optimize the experience of labour and childbirth for women and their babies. Based on the available evidence, a new labour care guide (LCG; Figure 1) was designed as a “next-generation” partograph to help health care providers (HCP) effectively monitor the well-being of women and babies during labour through regular assessments and reporting. Unlike the traditional partograph, the new LCG tool aims to stimulate shared decision-making between HCPs and women in order to promote women-centered care. The new LCG also aims to continuously remind practitioners to offer supportive care throughout labour and childbirth, and remind them of what observations need to be regularly made during labour to identify any emerging deviations from normal or complications in mother and/or baby. According to WHO, this LCG further provides new reference thresholds and alerts for abnormal labour observations which are meant to trigger specific actions. In adding all these components, the tool is expected to support prompt audits and minimize over-diagnosis and under-diagnosis of abnormal labour events and the unnecessary use of interventions such as caesarean sections and or labour augmentation, and thus improved quality of labour care and childbirth.

Identifying and scaling up such context-specific interventions have great potential to improve quality of healthcare and outcomes in pregnancy, childbirth and immediate postnatal period [13–16]. The Labour Care Guide manual has been developed by WHO to help skilled health personnel to successfully use the tool. However, the tool has not been adapted and adopted to be used as a decision-making tool in any low- and middle-income country maternity care settings, with correspondingly high rates of prolonged/obstructed labor, unnecessary interventions, maternal and perinatal mortality and morbidity. In this study, we aimed at customizing, characterizing and refining the new WHO labour care guide to suite health care provider needs in monitoring labour in South Western Uganda using iterative development methodology

## Methods

### Study design

We aimed at iteratively developing, customizing, contextualizing and refining a user-centered tool that would be comprehensive and easy-to-use, so as to optimize outcomes and long-term use by HCPs while monitoring labour in Uganda (Figure 2). The following steps were followed. We carried out stakeholder interviews to identify facilitators of continuous labour monitoring in Uganda, current user needs, and perceived LCG tool complications or challenges. This feedback informed the first modification of the WHO LCG into prototype one. We conducted HCP exit interviews following use of prototype one for at least 2 months to identify further needs. The suggested modifications resulted into prototype two. After deployment of this tool (prototype 2) for further use by HCPs in Mbarara city, we conducted two Focus Group Discussions (FGDs) involving midwives, interns, residents and consultants to identify any further user needs. These were integrated into the LCG tool to make prototype three. We then conducted the first exit expert panel interviews of 10 HCPs to further refine components of LCG prototype three. Prototype three was deployed for use/pilot test at MRRH for at least 2 months. The final feedback was obtained following a comprehensive presentation and review from two expert conferences, the 3^rd^ National safe motherhood conference (NSC2023), and the 18^th^ Association of Obstetricians and Gynecologists of Uganda to refine and consolidate all the proposed modifications into the final prototype for Uganda, ready for evaluation (Figure 3a and b). These steps were aimed at refining and customizing a user-centered tool for easy uptake and use within an established intervention development framework [17–19]. Data collection took place between 1st of July 2023 and 30^th^ November 2023.

### Study setting

Mbarara city just like elsewhere in Uganda has a public health system that is organized in five tiers, with a regional referral hospital and four levels of community health centers. Staffing and available maternity services vary across the four levels: HCIII also referred to as basic emergency obstetric and newborn care (BeMONC) facilities carry out vaginal deliveries, whereas HCI and HCII serve as low resource referral units. HCIVs and hospitals, referred to as comprehensive emergency obstetric and newborn care (CeMONC) facilities conduct caesarean deliveries and offer blood transfusion services [1]. Several private facilities operate in parallel to the public health system to provide maternal healthcare. There is a total of five HCIII, one HCIV and one regional referral hospital (MRRH) that serves the Southwestern region, where most of deliveries are high-risk [20]. MRRH also serves as the teaching hospital for Mbarara University of Science and Technology (MUST), whose Obstetrics and Gynaecology department has 14 Obstetricians and Gynaecologists, 31 Residents, 10 Intern doctors, and 38 Midwives. The nurse-to-patient ratio stands at about 1:25 [21]. The department performs about 10,000 deliveries annually, majority of whom deliver vaginally[22]. Women who deliver from the department are usually transferred from the antenatal clinic, referred from peripheral health units, or come directly from the community. Although often incompletely implemented, all mothers in labour are ideally monitored using a partogram-a graph of labour parameters and cervical dilation over time with pre-printed alert and action lines designed to prompt intervention if a woman’s curve deviates from the expected course. Additionally, despite decades of HCP training, rates of partograph completion and utilization to make critical decisions remain sub optimal in Mbarara district, Uganda and similar settings, with correspondingly high maternal and neonatal/still birth mortalities [23–26]. The fetal heart rate in Uganda is monitored manually per clinician judgment using the Pinard. Only one electronic monitor (CTG machine) and one ultrasound scan are available at MRRH, and are occasionally used to confirm presence or absence of regular cardiac pulsations, but not for monitoring labour [27]. Mbarara District is located ∼270 kilometres Southwest of the capital, Kampala [28].

#### Prototype development and pilot testing

We utilized a Behavioral Change Taxonomy [29, 30] to identify, refine/modify and characterize key LCG components needed to optimize knowledge of danger signs, cues, prompts, timely plan action, and maximize its long-term use and impact among HCP LCG users. We also used credible source of information from the intended HCP users, plus experts to customize information and instructions needed for self-monitoring, perform the needed behavior (complete LCG during monitoring) and improve HCP interaction for support and problem solving.

### Stakeholder interviews

We enrolled and interviewed 30 HCPs and MOH/WHO officials between June and September 2023 to achieve saturation of data. HCPs were purposively selected from 4 facilities across Mbarara city, Southwestern Uganda to represent diverse labour monitoring experiences and ideas about the new WHO LCG. We also explored challenges and opportunities of labour monitoring in the present times. We included HCPs and MoH/WHO officials exposed to the new WHO LCG. The data from these interviews was used to identify key LCG components and characterize/modify and customize the new WHO LCG into a useable version (prototype 1) for HCPs in Southwestern Uganda. The detailed data from these stakeholder interviews has been published elsewhere [31].

#### HCP exit Interviews

A total of 10 exit HCPs interviews were done following use of the modified LCG (prototype 1) for at least 2 months. Participants of different cadre and qualifications were screened and purposively selected from Mbarara city health facilities to explore user experiences, opportunities and ideas to improve the prototype. This data was synthesized and used to further modify the LCG. This modified version (prototype 2) was deployed in two Mbarara city facilities for testing. All interviews were open-ended and covered a wide range of topics following the interview guide developed to explore ease of use, perceived usefulness, the HCP attitude to LCG use, preference, effort and performance expectancy, as well as other useful topics like self-efficacy, behavioral intention to use and actual use of the modified WHO LCG. A brief data on demographic information (e.g., age, qualification, work experience) was collected. All interviews were conducted in a private location agreed upon by the HCPs. All interviews lasted 40-60 minutes, and written informed consent was obtained from all participants at the start of each interview session. All interviews were recorded digitally, with participant’s permission and transcribed verbatim.

#### Focus Group Discussions

Two FGDs, each containing 10 HCPs of all cadres that were exposed to prototype 2 were conducted to explore experiences and feedback on the modified prototype 2. This input/feedback was synthesized, and ranked to modify/refine components of a subsequent prototype 3.

### Expert Pannels

This prototype 3 was further subjected to two expert panels each comprising of 10 HCPs of different cadres (2 certificate midwives, 8 diploma midwives, 6 bachelor’s degree nurses and residents, and 4 consultant obstetricians) to provide feedback on specific LCG prototype 3 components, plus experiences of this version’s functionality and ease-of-use to monitor labour in Southwestern Uganda. Following these two expert panels, we further refined some components of the modified LCG to produce the final LCG prototype for Uganda, ready for pilot testing.

#### Final LCG prototype testing

The refined prototype 3 was deployed for use and pilot tested for acceptability and appropriateness among HCPs conducting deliveries at Mbarara Regional Referral Hospital (MRRH), the busiest facility in Southwestern Uganda for at least two months. We used a structured questionnaire to obtain demographic information, as well as feedback on appeal, ease of use, complexity, content, usefulness, tool’s preference, time taken to plot, fill and complete tool after assessment, its appropriateness and feasibility in this setting.

Prototype 3 was presented for final feedback and review from two expert conferences; the 3^rd^ National safe motherhood conference (NSC2023), and the 18^th^ Association of Obstetricians and Gynecologists of Uganda. The presentation and engagement with key stakeholders at these two expert conferences was aimed at obtaining direct feedback towards refining and consolidating the proposed modifications into the final prototype for Uganda, ready for evaluation (Figure 3a and b). This final prototype will be evaluated in effectively monitoring labour and improving clinical outcomes across Mbarara district in Southwestern Uganda as the next step.

## Data Analysis

We described demographic and clinical data for all interviewed participants using standard descriptive statistics. Qualitative analysis included repeated review of transcripts to identify relevant experiences, ideas and preferences on labour monitoring and LCG use. We coded qualitative data using NVivo (version 12.0; Melbourne, Australia), and iteratively reviewed and sorted codes to identify repeated themes using inductive content analysis [32]. We used illustrative quotes taken from the qualitative interviews to illustrate the context and meaning in each theme. Data analysis was done jointly by GRM and WT to ensure consistency in coding.

### Ethical considerations

Ethical clearance was obtained from the Faculty Research Committee in the Faculty of Medicine and the Research Ethics Committee (REC) at Mbarara University of Science and Technology (MUST-2023-808). Study site administrative Clearance was obtained from the Hospital Director of Mbarara Regional Referral Hospital, Mbarara District Health officer for Mbarara District and the City Health Officer for Mbarara City. We sought approval from the National Council for Science and Technology in Uganda (UNCST) – HS2864ES

### Ethical Compliance with Human Study

This study was conducted in compliance with the ethical standards of the responsible institution on human subjects as well as with the Helsinki Declaration

## Results

We interviewed a total of 120 participants for this development phase. The median age of the interviewed HCPs was 36 years (IQR;26-48), distributed across enrolled (14%) diploma (38%) midwives, bachelors/Residents (31%) and consultants (18%). The mean years of practice for the HCPs was 11.8 (SD=4.6), and all HCPs had at least 12 months of exposure to the LCG (Table 1).

**Table1:** Demographic characteristics of study participants.

| Characteristic | Stakeholder<br>Interviews<br>(n=30) | HCP<br>interviews<br>(n=10) | FGD<br>Interviews<br>(n=20) | Expert<br>Panels<br>(n=20) | Pilot<br>(n=40) | testAll<br>Participants<br>(n=120) |
| --- | --- | --- | --- | --- | --- | --- |
| **Median age (IQR)** | 36 (27,54) | 38(28, 45) | 31(25,47) | 36(26,48) | 35(25,52) | 36(26,48) |
| **Mean duration of clinical practice in years (SD)** | 11.3 (7.7) | 15.2 (4.7) | 8(5.2) | 9.2(5.5) | 10.2(4.4) | 11.8(4.6) |
| **HCP Cadre, n (%)** |  |  |  |  |  |  |
| Certificate Midwife | 5(13.9) | 2(20) | 2(10) | 2(10) | 6(15) | 17(14.2) |
| Diploma Midwife | 10(27.8) | 2(20) | 8(40) | 8(40) | 17(42.5) | 45(37.5) |
| Bachelors/Residents | 10(36.1) | 4(40) | 6(30) | 6(30) | 11(27.5) | 37(30.8) |
| Consultant | 5(13.9) | 2(20) | 4(20) | 4(20) | 6(15) | 21(17.5) |
| **Mean time exposure to LCG (months)** | 12.3(4.2) | 12.0(3.8) | 13(2.5) | 12.4(2.5) | 12.0(2.5) | 12.3(2.8) |

### LCG prototype design and development

From all qualitative interviews, a number of modifications were made on the original WHO LCG tool (Figure 1), based on format and HCP’s perceived function/role/goal in monitoring labour and improving decision making or action planning. HCPs identified and defined prototype design and content needs that were expected to help them optimize LCG long-term use, and rates of completeness with its intended benefits. These modifications or customizations, also summarized in table 2, included; 1) Adjusting observation ordering and font size to facilitate an easy-to-use interface, flow, engagement and clarity; 2) Customizing LCG by adding key socio-demographic data compatible with existing programs to aid planning and managing risk; 3) Modification of key fields to suit local context; 4) Inclusion of key clinical notes and labour outcome data to facilitate auditing, accountability, reference, utilization and immediate postpartum care; 5) Presentation of LCG and protocols/guidelines in attractive colours, size, shape and content that match the theme of safe motherhood, facilitating familiarization, visual appeal, ongoing user support, awareness and education.

**Table 2:** Differences between the modified partograph, who LCG and the modified LCG.

|  |  |  |  |
| --- | --- | --- | --- |
| **Parameter** | **Modified WHO partograph** | **WHO Labour Care Guide** | **Modified Labour Care Guide Prototype 3** |
| Name of Health Center | No | No | Yes |
| In Patient Number | No | No | Yes |
| Name of Patient | Yes | Yes | Yes |
| Age | No | No | Yes |
| National Identification Number (NIN) | No | No | Yes |
| Woman's gravidity | Yes | No | No |
| Woman's Parity | No | Yes | Yes |
| LNMP | No | No | Yes |
| Weeks of gestation | No | No | Yes |
| Date and time of admission | No | Yes | Yes |
| Time of rupture of membranes | Yes | Yes | Yes |
| Type of labour onset (spontaneous or induced) | No | Yes | Yes |
| Medical and social risk factors | No | Yes | Yes |
| HIV/Syphilis and Hepatitis Test results | No | No | Yes |
| Supportive care interventions (labour companionship, pain relief, oral fluid intake, and maternal position) | No | Yes | Yes |
| Fetal heart rate (FHR) | Yes * | Yes** | Yes** |
| Presence of early, late, or variable decelerations | No | Yes | Yes |
| Normal lower limit of FHR | 110 | 110 | 120 |
| Amniotic fluid characteristics | Yes | Yes | Yes |
| Fetal position | No | Yes | Yes |
| Caput | No | Yes | Yes |
| Moulding | Yes | Yes | Yes |
| Woman's pulse and blood pressure (BP) | Yes* | Yes** | Yes** |
| Woman's temperature | Yes | Yes | Yes |
| Urine volume | Yes | No | No |
| Proteinuria and acetonuria | Yes | Yes | Yes |
| Duration and frequency of uterine contractions | Yes | Yes | Yes |
| Strength of contractions | Yes | No | No |
| Definition of active phase | Starting from 4 cm of cervical dilatation | Starting from 5 cm of cervical dilatation | Starting from 5 cm of cervical dilatation |
| Definition of "satisfactory" labour progress | Fixed 1 cm/hour time limit ("alert" and "action" lines) | Evidence-based time limits at each centimeter*** | Evidence-based time limits at each centimeter*** |
| Cervical dilatation | Yes | Yes | Yes |
| Descent of the fetal head | Yes | Yes | Yes |
| Values for "normal" | "Alert" and "action" lines for cervical dilatation; thick lines to identify parameters for normal FHR | "Reference threshold" values*** are listed for non-clinical and clinical parameters | "Reference threshold" values*** are listed for non-clinical and clinical parameters |
| Second stage section | No | Yes (all parameters except cervical dilatation) | Yes (all parameters except cervical dilatation) |
| Time when pushing begins | No | Yes | Yes |
| Identification of deviations from expected observations | No explicit way to document deviations from expected observations of any labour parameter, other than cervical dilatation to the right of "alert" and action lines and FHR 180 bpm or faster/100 bpm or slower | Requires circling any observations meeting the criteria in the "alert" column | Requires circling any observations meeting the criteria in the "alert" column |
| Assessment of findings | No | Yes | Yes |
| Plan of care | No | Yes | Yes |
| Provider's initials | No | Yes | Yes |
| Record of labour outcomes and other important labour and post-partum observations and management on the flip side of the paper | No | No | Yes |
| *Values are plotted on a graph<br>**Values are written in the appropriate cell<br>***Reference threshold values for labour observations define normal, expected ranges for the different parameters. They are intended to trigger reflection and specific action(s) if an abnormal observation is identified. |  |  |  |

#### 1) Adjusting observation ordering and font size to facilitate an easy-to-use interface

According to HCP interviews, adjusting the font size would make instructions, labour observations and foot notes easy to read and follow. All HCPs also suggested that the LCG sequence of observations needed to be re-aligned to be in synch with the flow of activities and sequence of normal labour process. For example, 1) Assessment section of the shared decision making was modified to include <u>impression/ assessment.</u> The impression was HCP’s preferred reference to diagnosis following examination and clinical assessment as commonly interpreted locally, 2) Under shared decision making, “plan” was suggested to be modified to “action plan” to prompt the attending clinician to make or document a suitable action plan, following complete assessment and interpretation of all labour observations, including alerts, 3) Section 6 (Medication) was suggested to come after shared decision-making (originally as section 7). This was thought that the medications would be prescribed following complete assessment and documentation of action plan preceding the assessment/ impression or diagnosis. All these modifications (as seen in Figure 4) were suggested by HCPs to have potential to improve the tool’s user interface, facilitate logic flow of information, HCP engagement and clarity during labour monitoring. One of the midwives who has had over 10 years’ experience in managing labour said,

> “You see, we work in busy units where time is of essence. It is very helpful to arrange these observations in a manner that makes sense for most of us, but also in a way that helps us to quickly document and make sense of these observations easily and plan with others”.

#### 2) Customizing LCG by adding key socio-demographic data compatible with existing programs

HCPs noted that there were key socio-demographic data that was missing in the original WHO LCG that needed to be added in order to aid planning care and managing risk among women enrolled in labour. These included; 1) the Hospital/facility name, in-patient number (IPN), patient age and National Identification number (NIN), for easy patient identification and inter-facility tracking in times of patient transfer/referral. These identifications are currently routine in all public facilities in Uganda; 2) Last normal menstrual period (LNMP), and weeks of gestation, and specific dates and time format of other events like labour onset, active labour diagnosis and raptured membranes. These would improve clarity, consistency and uniformity in documenting labour events and risk assessment; 3) The HCPs noted data that were already part of MOH’s existing routine priority labour monitoring parameters/program that guide any HCPs to identify and manage/plan women at risk of mother-to-child infection transmission. These included prevention of mother to child transmission (PMTCT) code, Syphilis and Hepatitis B test results, aimed at screening for HIV/AIDS, syphilis and Hepatitis B respectively for all mothers in labour (also known as triple elimination). This was suggested to make LCG compatible with the existing programs, and makes planning and management of risk easier and better when included on the same form. According to a Senior Health Officer, with 12 years of experience in labour monitoring,

> “Each mother is unique and so is their management plan. A lot of this information gives the managing team a good perspective on the nature of risk or mother you are managing. It is a good way to quickly translate information to another team taking over for example, the women who are at risk of infection such as HIV and others as per the existing MOH policies”.

#### 3) Modification of key fields to suit local context

HCPs interviewed during expert panel interviews noted that the lower limit of the baseline fetal heart rate needed to be modified from ≤110 to ≤120 beats per minute, as previously captured on the old partogram. This was suggested to provide a safety margin in case of routine patient transfer/referral from the basic to the comprehensive emergency obstetric and neonatal care units that are usually far apart with limited transportation/ambulance system. However, in an attempt to create space, some HCPs argued that fetal heart rate deceleration record was redundant and not applicable for most public facilities since it required CTG machines that are unavailable, and not mandatory in most Ugandan basic and comprehensive emergency obstetric care facilities, except for some regional referral and private hospitals. This section was maintained following expert panel review for the benefit of the facilities that had the capacity to acquire them and for future plans and developments in obstetric care and management.

The oxytocin row under the medication section was modified to accommodate the oxytocin dose in International Units (IU) administered in a 500mL unit of available crystalloids (normal saline or ringers’ lactate) locally in Uganda. The 1 litre unit provided for in the WHO LCG is not available on market in Uganda. An additional row to accommodate the rate of oxytocin administered at a particular time in drops per minute was added to aid detailed documentation and interpretation of augmented labour progress. The hourly boxes were also bisected to accommodate the dose and rate for every 30 minutes of incremental oxytocin administration. Similar columns and rows were added to accommodate the type and volume of intravenous fluid (IV fluids) used in labour. The separation would make the section easy to record the different fluids and volume during labour at a particular time, making it less squeezed in one small box provided by the WHO LCG.

The row for capturing other medications, other than oxytocin used during active labour was deemed redundant during active labour monitoring. Any other medicines (not labour care monitoring) including those given for co-existing infection, severe preeclampsia/eclampsia, malaria, epidural, Antiretrovirals (ARVs), analgesics, etc were recommended to be removed from the active labour monitoring section six, to be recorded on the reverse side of the modified LCG to include all other medications, and tranexamic acid, oxytocin/other uterotonics given during active management of third stage of labour. A senior midwife aged 50-55 year argued,

> “The LCG indicates the 1 litre intravenous bottles, which are not available in Uganda and so this recommendation would be redundant and not appropriate here. Secondly, the medications other than oxytocin that is used during labour monitoring are not usually part of labour monitoring and could be removed from the chart and recorded elsewhere to avoid confusion. I also noted that the basic maternity service centres cannot do C-sections, and often times need a good transfer window to transfer women with foetal heart of at least less than 120 beats per minute. Moreover, I think in our setting with the kind of poor ambulance system and roads, many women may take long to transfer and so that window would really be useful and appropriate for us here”.

#### 4) Inclusion of key clinical notes and labour outcome data

Data indicated that the WHO LCG needed to be more inclusive, and comprehensive. HCPs suggested inclusion of vital clinical and labour outcomes on the LCG’s reverse side to facilitate easy/ quick auditing, accountability, reference and immediate postpartum care. According to the HCPs, inclusion of this information on two pages of one A4 paper would mitigate the extra cost on stationary that could pose a major challenge especially at basic emergency obstetric and new born care centers. The comprehensiveness of data recorded on this modified A4 paper was also thought to make workload easier if LCG was comprehensive enough to be considered to have information enough to make an accurate, appropriate and prompt decision during and after active labour monitoring. HCPs noted that with these considerations, LCG had potential to be the only required labour monitoring and management tool on ward, avoiding the challenge of what was termed as over-documentation in different places, including patient files, notes, treatment forms, fluid balance charts, temperature charts, something they hoped would motivate and encourage tool completion and utilization. Notably, many of these outcomes are a part of the Ugandan partogram that is poised to be replaced; a few have been proposed to prompt action based on recent data indicating the status of women that show up at facility in active labour for the first time. These modifications include; 1) space for writing number of antenatal care visits and date of first antenatal care visit, 2) space for prompting HCP to investigate or add the last hemoglobin level taken during ANC or labour, 3) space for any additional clinical notes not captured within the 7 sections of the LCG, e.g. use of herbal medicine during labour, disability, abnormalities, other non-labour-related diagnoses or assessments and observations, 4) outcomes of labour, e.g. date, time and type of delivery, duration of first and second stages of labour, amount of blood loss, placental examination, episiotomy/tears and repairs given if any, baby outcomes e.g., status (alive/dead), Apgar score, sex, birth weight, abnormalities, medicines given to baby including tetracycline, vitamin K, nevirapine syrup for HIV exposed newborns, 5) space for recording the name and cadre for the HCP that conducted the delivery and their assistant, 6) space for documenting immediate postpartum care given, record for postdelivery blood pressure, pulse, temperature, initiation of breastfeeding within 1 hour, new born temperature and immunizations given prior to discharge (see details in Figure 4). All these additions were suggested to make the modified LCG for Uganda a “one stop, one A4-size paper reference tool” that was comprehensive to make labour monitoring and management easier and effective. According to a 46 −50-year-old senior midwife, who doubles as a maternity unit in charge,

> “The clinical outcomes and key notes can be added on the flip side of this LCG to cut back on stationery and over filling and recording the same things in different places. We can then have everything we need in one place, which can trigger a quick plan for action by just running through one form…This makes life easier and workload lighter without using more stationery and forms that is often costly and unavailable”.

#### 5) Presentation of LCG and its protocols/guidelines in attractive colors, size, shape and content that match the safe motherhood theme

HCPs noted that change generally, was always difficult for people to appreciate and completely embrace new tools and interventions after long-term use of previous partogram. They suggested presentation of the new LCG and its protocols/guidelines in attractive colors, size, shape, content and format, with a good visual appeal that would facilitate both champions and resistors familiarization, interaction, ongoing user support, awareness and education. According to a 26–30-year-old midwife with 5-10 years of experience,

> “Being in a different and exclusive colour from the old partograph can easily capture anyone’s attention, awareness and to be reminded about its use and why it is there”.

## Pilot testing

### HCP Interviews and FGDs

All HCP involved in the exit interviews, and two FGDs seemed to like the modified WHO LCG prototype. HCPs described the prototype as simple, easy-to-use, and quick to fill/plot compared to the old partograph. HCPs who reported prior training and exposure for at least one months seemed to exhibit high enthusiasm to complete and correctly use the LCG. Assessment of acceptability; perceived use, usefulness, attitude and behavioral intention to use the modified WHO LCG showed that all the HCPs liked the current design and described it as detailed, comprehensive and customized enough to meet HCP user needs, as the tool captured all vital parameters on a one-A4 pager, with potential to reduce over documentation, save time and reduce workload. HCPs also presented this design as having potential to not only allow a one stop quick reference for improved clinical decisions but also could promote interaction, responsibility, accountability, team work and confidence among HCPs, women and others on the care team. Largely enthusiastic HCPs presented this modified LCG as accurate, dependable, and satisfying, and that its parameters were largely familiar, thus requiring minimal additional training to use effectively. The narratives in these interviews seemed to resonate with the ones reported above, and previously reported elsewhere [31]. The great enthusiasm expressed by all HCPs also seemed to present new opportunities to actually use the tool long-term to improve intrapartum care for women in labour as summarized in Figure 4 below.

### Pilot testing the Final LCG prototype

Pilot testing the refined and final prototype followed feedback from FGDs and expert panelists. A total of 40 HCPs from MRRH approved and liked the new and modified LCG, finding it appealing, welcome, easy to use and useful to themselves and other HCPs (Table 3). All HCPs also liked the new LCG and would recommend it to others for use in labour monitoring. All interviewed HCPs also found it appropriate and implementable. All HCPs preferred the new LCG for recording labour progress and majority took less than 2 minutes to completely record, plot or fill observations on the LCG after each labour assessment.

**Table 3:**
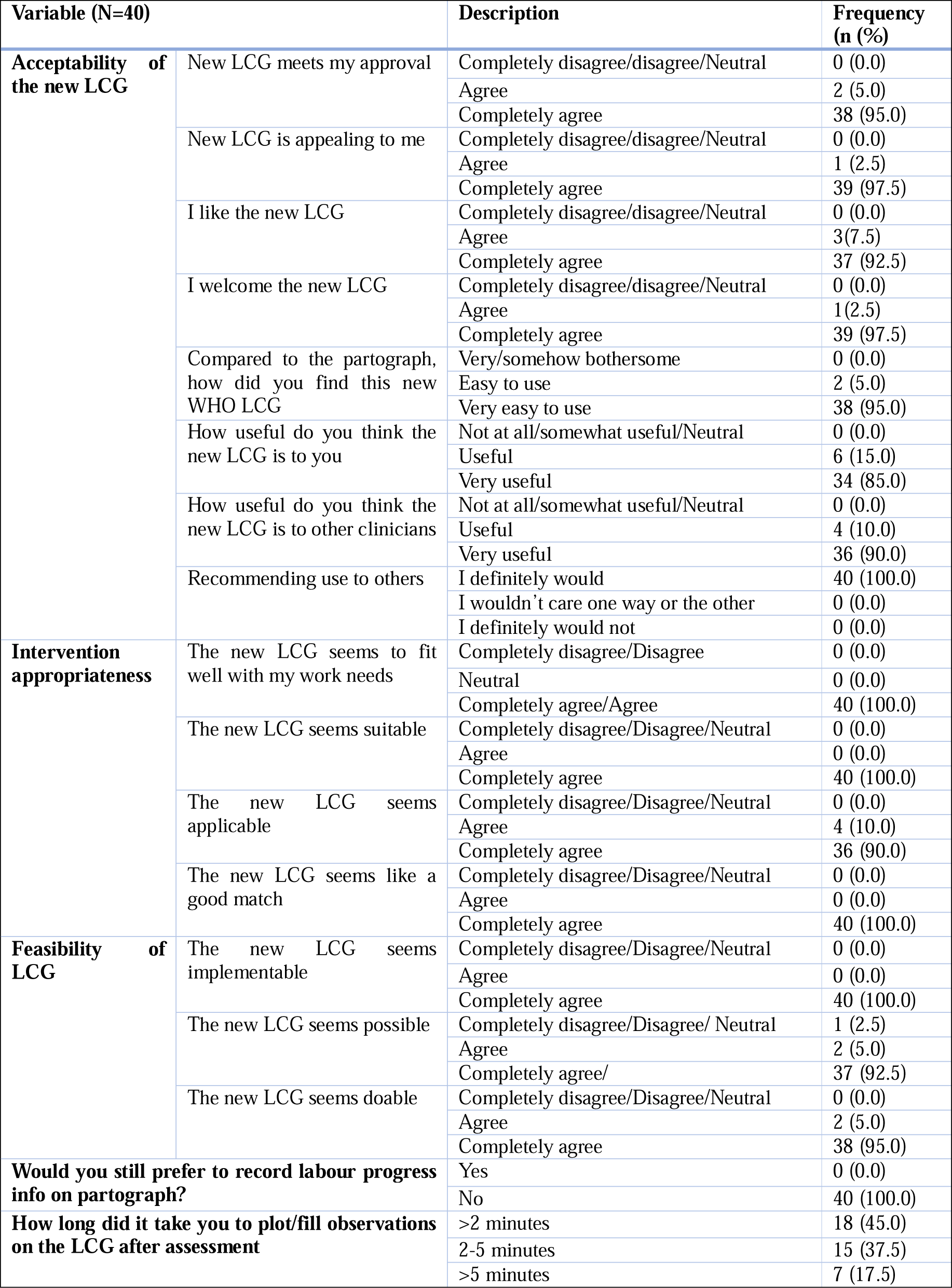
Acceptability of the modified LCG prototype 3.

## DISCUSSION

The enhancement of better quality, evidence based and respectful care during labour and childbirth requires concerted efforts towards better maternal and child health outcomes. This study sought to utilize iterative approaches to customize and modify the new WHO Labour Care Guide to develop a locally contextualized acceptable tool that is applicable within Ugandan and other similar settings, prevalence in LMIC to enhance ease of use, easy uptake and sustained utilization to maximize impact [17–19]. The major modifications on the new WHO LCG to the “modified Labour Care Guide prototype” for Uganda included; a) adding key socio-demographic data for better planning and risk management, b) increasing the font size and labour observation re-ordering to facilitate an easy-to-use interface, flow, engagement and clarity c) inclusion of key cues to action such as triple elimination code, Hb level and others, as well as additional clinical notes and labour outcome data for auditing, accountability, reference, utilization and immediate postpartum care, d) modification of key fields to suit local context, for example, modifications to accommodate the oxytocin dose in International Units (IU) administered in a 500mL unit of available crystalloids (normal saline or ringers’ lactate) locally in Uganda, including the lower limit of the normal fetal heart rate from 110 to 120 beats per minute to provide a safety margin for referral since 55% of deliveries occurred at BeMONC sites (HCIIs and IIIs) unable to conduct caesarean section when required[33], e) making the guidelines visually appealing in terms of colour, shape and size to match safe motherhood the themes.

Findings from this study show that HCPs approved and liked the modified WHO LCG for Uganda because of its potential ease-of-use and ability to record all labour parameters on one A4 size paper, and indicated they would recommend it to other clinicians for use in labour monitoring. They also reported that the modified WHO LCG was appropriate and implementable, took shorter time of less than 2 minutes to complete plotting and filling labour observations after each assessment. This is in tandem with findings reported in North India where the time of active labour was significantly shorter in the intervention group compared to the control group although both groups had similar outcomes in terms of maternal complication, hospital stay and APGAR score [34].

The utilization of user-centered approaches has potential to facilitate optimum intervention exposure, delivery, hands-on-skilling, responsiveness and ability to use the intervention to effectively monitor labour, and make key decisions regarding labor interventions to improve outcomes. This is aimed at behavioral change regarding the intervention usage in terms of awareness, accurate delivery reporting, comprehension and decision making [17]. The goal of this approach was to make the modified labour care guide easy to use, inclusive, educative, supportive, with periodic engaging prompts and cues to aid action in real time [19]. Our findings demonstrate that the involvement of clinicians in developing interventions gives them a voice and a sense of ownership thus making the modified WHO LCG’s acceptability and uptake easier. This is in agreement with a study by Skivington and colleagues, that noted that the development of interventions always involves iterative cycles that includes obtaining feedback from stakeholders, innovation implementation and assessing the acceptability of these interventions and starting the cycle again until all the iterations produce the required changes [35]. Therefore, it is important for implementers to introduce interventions for use after carefully considering the context within which the intervention will be implemented to facilitate sustained integration, use and better health outcomes. Indeed, some scholars have shown that end-user designs that utilize iterative approaches in characterizing, adopting and implementing these new approaches improve intervention uptake and sustained use among intended users [7].

We modified/refined the new WHO LCG to include all the needed parameters for labour care monitoring in Uganda and improve maternal-foetal clinical and labour outcomes. The refined prototype aimed at facilitating timely auditing, accountability, reference, planning and immediate postpartum care without making it complicated to fill. Given the high patient doctor ratio in low resource settings like Uganda where the number of patients is higher compared to the available obstetric care providers which increases workload [36], such an intervention that takes a shorter time to fill could potentially enhance effective labour monitoring and better health outcomes. Indeed, many scholars [37–43] have shown that one of the challenges that hinder the utilization of the partograph is its complexity, subjectivity, which take a lot of time to fill and interpret alongside other required and parallel patient records amidst shortage of staff in health facilities that unnecessarily increases the workload of the already overwhelmed HCPs. The ability of the refined LCG prototype to be more inclusive, comprehensive and easy to use makes it a one stop user friendly tool that could facilitate easy/ quick auditing, accountability, reference to make accurate, appropriate and prompt decision during and after active labour monitoring, while mitigating the extra cost on stationary, over filing, and over documentation that could pose a major challenge especially at basic emergency obstetric and new born care centers with limited workforce and stationery. This could enable HCPs take considerably shorter time to fill out and facilitate prompt action during labour management.

Our study attempted to customize and tailor this promising labour monitoring tool to fit within the local settings and enhance usability and acceptability. The involvement of end users to solicit their ideas, user perspectives, experiences and feedback during the development of this new tool was very crucial in ensuring that the final prototype meets their expectations and improve their overall user experience. These systematically participatory design approaches have been observed to make end users feel that their voices are heard and that their concerns have been catered for during the design, thus ultimately improving uptake and health outcomes [44]. Other studies have shown that interventions that are simply replicated in other settings may yield suboptimal outcomes compared to those locally contextualized and adapted to attain a good fit between intervention and context [45]. Therefore, characterization and refining the WHO LCG may save unnecessary time and money by addressing the potential user challenges, needs, preferences and issues early on to build trust, improve acceptability, uptake, scale up and integration in routine care by the HCPs in Uganda and similar settings.

Our study presents a number of strengths. It is nested in the principles of participatory design that emphasizes the involvement of key end user stakeholders in intervention development of a user-friendly, ccontext-specific tool aimed at supporting HCPs to effectively monitor labour in Southwestern Uganda. This approach could potentially facilitate acceptability, functionality, usability as well as reinforce ownership, inclusiveness, comprehensiveness, engagement and uptake among the targeted end user HCPs in Uganda and similar settings, subject to routine challenges of stationery and limited health work force. Secondly, this is the first study that reports the customization, tailoring and contextualization of the new WHO labour care guide for Uganda and similar settings, and grounded in evidence-based conceptual frameworks, making our findings meaningful, generalizable and grounded. Our study also recognises some limitations. We did not evaluate the feasibility, rate of completeness, incremental cost, sustainability and effectiveness of this new prototype to improve maternal-foetal clinical and labour outcomes. Evaluation of our final prototype is ongoing to document effectiveness and its diagnostic validity compared to the partograph.

### Conclusion

Our study describes an iterative process for customizing, tailoring, refining and pilot testing the novel WHO LCG tool for labour monitoring among HCPs end users in southwestern Uganda. Involvement of targeted end-users in modifying/refining the WHO LCG tool was observed to improve acceptability, ownership, inclusiveness, comprehensiveness, engagement and uptake. The modified LCG for Uganda was found to be simple, appropriate, easy-to-use, with all HCPs preferring the new LCG for recording labour parameters and labour outcomes on the reverse page, compared to the partograph. It was found quick to fill/plot with majority taking less than 2 minutes to completely record all observations after each labour assessment. It was described as detailed, comprehensive and customized to meet local context HCP user needs, capturing all vital labour parameters on a one-A4 pager, with potential to promote HCP-labour companion-patient interaction, responsibility, accountability, team work and confidence among HCPs, women and others on the care team. Further research to evaluate the modified LCG prototype’s large-scale feasibility, acceptability and effectiveness in helping HCPs timely detect deviations from normal labour and prevent complications compared to the traditional partograph in different facilities and or settings is warranted.

## Informed Consent

We obtained written informed consent from all study participants before data collection started.

## Data Availability

All data produced in the present study are available upon reasonable request to the authors

## Acknowledgement

The authors acknowledge all health Care Providers who participated in all listed steps followed in development and pilot testing of the WHO Labour Care Guide into the Modified Labour Care \guide Prototype presented in this manuscript

## Conflict of Interest

All the authors declare no conflict of interest.

## Financial Disclosure or Funding

None

## Authors Contribution

MRG developed the research concept, supervised data collection, analyzed data alongside WT, ACE, and wrote the first draft of this manuscript under the supervision and mentorship of BKJ, FTY, ECA and TW. All authors reviewed and approved the manuscript

## Availability of data and materials

A couple of de-identified transcripts have been provided alongside this submission. The parent study is still ongoing and appropriate data sets will be available on request from the authors. Eventual data sets will be deposited in a repository for future reference.

## List of Abbreviations

ANC: Antenatal Care
BeMONC: Basic Emergency Obstetric and Newborn Care
CeMONC: Comprehensive Emergency Obstetric and Newborn Care
CFIR: Consolidated Framework of Implementation Research
HC: Health Centre
HCP: Heath Care Provider
IPN: In-patient Number
LCG: Labour Care Guide
LMIC: Low- and Middle-Income Countries
LNMP: Last normal menstrual period
MMR: Maternal Mortality Ratio
MOH: Ministry of Health
MRRH: Mbarara Regional Referral Hospital
MUST: Mbarara University of Science and Technology
NIN: National Identification number
PMTCT: Prevention of Mother To Child Transmission () code
REC: Research Ethics Committee
UDHS: Uganda Demographic Health Survey
UNCST: Uganda National Council for Science and Technology
WHO: World Health Organization

## Notes

### Competing Interest Statement

The authors have declared no competing interest.

### Clinical Trial

NCT05979194

### Funding Statement

This study did not receive any funding

### Author Declarations

Ethics committee of Mbarara University of Science and Technology approved this work

